# Creating a safe workplace by universal testing of SARS-CoV-2 infection in patients and healthcare workers in the electrophysiology unit having no symptoms of COVID-19: a multi-center experience

**DOI:** 10.1101/2020.07.14.20153494

**Authors:** Sanghamitra Mohanty, Dhanunjaya Lakkireddy, Chintan Trivedi, Bryan MacDonald, Angel Mayedo, Domenico G Della Rocca, Rakesh Gopinathannair, G J Gallinghouse, Luigi Di Biase, Rodney Horton, Robert Canby, Andrea Natale

## Abstract

**Background:** As the coronavirus cases continue to surge, the urgent need for universal testing to identify positive cases for effective containment of this highly contagious pandemic has become the center of attention worldwide. However, in spite of extensive discussions, very few places have even attempted to implement it. We evaluated the efficacy of widespread testing in creating a safe workplace in our healthcare community including staff and patients.

Furthermore, we assessed the rate of new infections in patients undergoing electrophysiology (EP) procedure, to see if identification and exclusion of positive cases facilitated establishment of a risk-free operating environment.

**Methods:** Universal testing was conducted in subjects with no symptoms of COVID-19 including patients and their caregivers and staff in our electrophysiology units along with the Emergency Medical Service (EMS) staff (n=1670)

**Results:** Of 1670, 758 (45.4%) were EP patients, and the remaining 912 were caregivers, EMS staff and hospital staff from EP clinic and lab. Viral-RNA test revealed 64 (3.8%) positives in the population. A significant increase in the rate of positives was observed from April to June, 2020 (p=0.02). Procedures of positive cases (n=31) were postponed until they tested negative at retesting on day 14. Staff testing positive (n=33) were retested before going back to work after 2 weeks. Because of suspected exposure, 67 staff were retested and source was traced. No new infections were reported in patients during the hospital stay or within 2-weeks after the procedure.

**Conclusion:** Universal testing to identify positive cases was helpful in creating and maintaining a safe working environment without exposing patients and staff to new infections in the EP unit at the hospital.

**Trial Registration Number:** clinicaltrials.gov: NCT04352764

## Introduction

SARSCoV-2 infection causing Coronavirus Disease 2019 (COVID-19) became a global pandemic in a very short period of time affecting humanity in an unprecedented way (1). Since then, several million patients have been infected and hundreds of thousands have died in the United States alone. High infection rate and mortality coupled with limited knowledge about the SARS-CoV-2 virus have made the management of this pandemic extremely challenging. Subsequently, social distancing and lock down measures have been put in place to control the spread. These drastic yet important measures have not only affected the economy and social life, but also have dramatically impacted essential services including cancellation of all elective procedures in hospitals. Furthermore, this pandemic has left millions of patients in a state of panic and compromising access to care. For the fear of contracting the highly contagious COVID-19 infection, many ill patients sequestered at home have been reported to be presenting in later stages of their disease progression leading to poor outcomes and mortality (2).

A rational tactic to control this rapidly spreading infectious disease would be by identification of positive cases using universal testing followed by contact-tracking and isolation of confirmed cases. Consequently, this would not only curtail the new infection rates in the community, but also would be helpful in titrating restrictive measures instead of imposing sweeping lockdowns on all. Although this approach seems absolutely logical, testing in the USA and world-wide has been limited to symptomatic subjects only and universal testing has been implemented in a very small number of communities (3-5). We designed a model of mass-testing for the electrophysiology service line in an attempt to create a COVID-safe environment and keep providing the much needed procedural care. Here, we report the impact of universal testing on lowering the risk of transmission of SARS-CoV-2 infection in the hospital setting among the patients and healthcare workers in our hospitals.

## Methods

In this consecutive series, patients scheduled for electrophysiology (EP) procedures and their caregivers as well as staff in the EP subspecialty with no symptoms of COVID-19, were tested for the SARS-CoV-2 viral-RNA and serology tests. The aim was to screen for positive cases in asymptomatic staff and patients in population presenting to the EP service. The serological test was performed utilizing COVID-19 Rapid Test (Premier Biotech Labs, 723 Kasota Avenue SE, Minneapolis, MN 55414 and Confirm Biosciences, San Diego, CA) that uses a drop of blood from finger-prick to detect presence of IgM and IgG in 15 minutes or less. Nasopharyngeal swab was tested for viral RNA using ID NOW test kit from Abbott Diagnostics Scarborough, Inc., Maine. All test kits were cross verified internally by testing them against known COVID positive and negative patients.

Participants were included only if they were asymptomatic. They were screened based on a survey that assessed their travel and contact history along with symptoms. Pre-existing medical conditions were also noted during the screening procedure.

Exclusion criteria included body temperature higher than 38^°^C, symptoms of COVID-19 and history of close contact with a known infected patient in the last two weeks. These patients were triaged to viral-RNA test using nasal swab (FIGURE 1).

**Figure 1:**
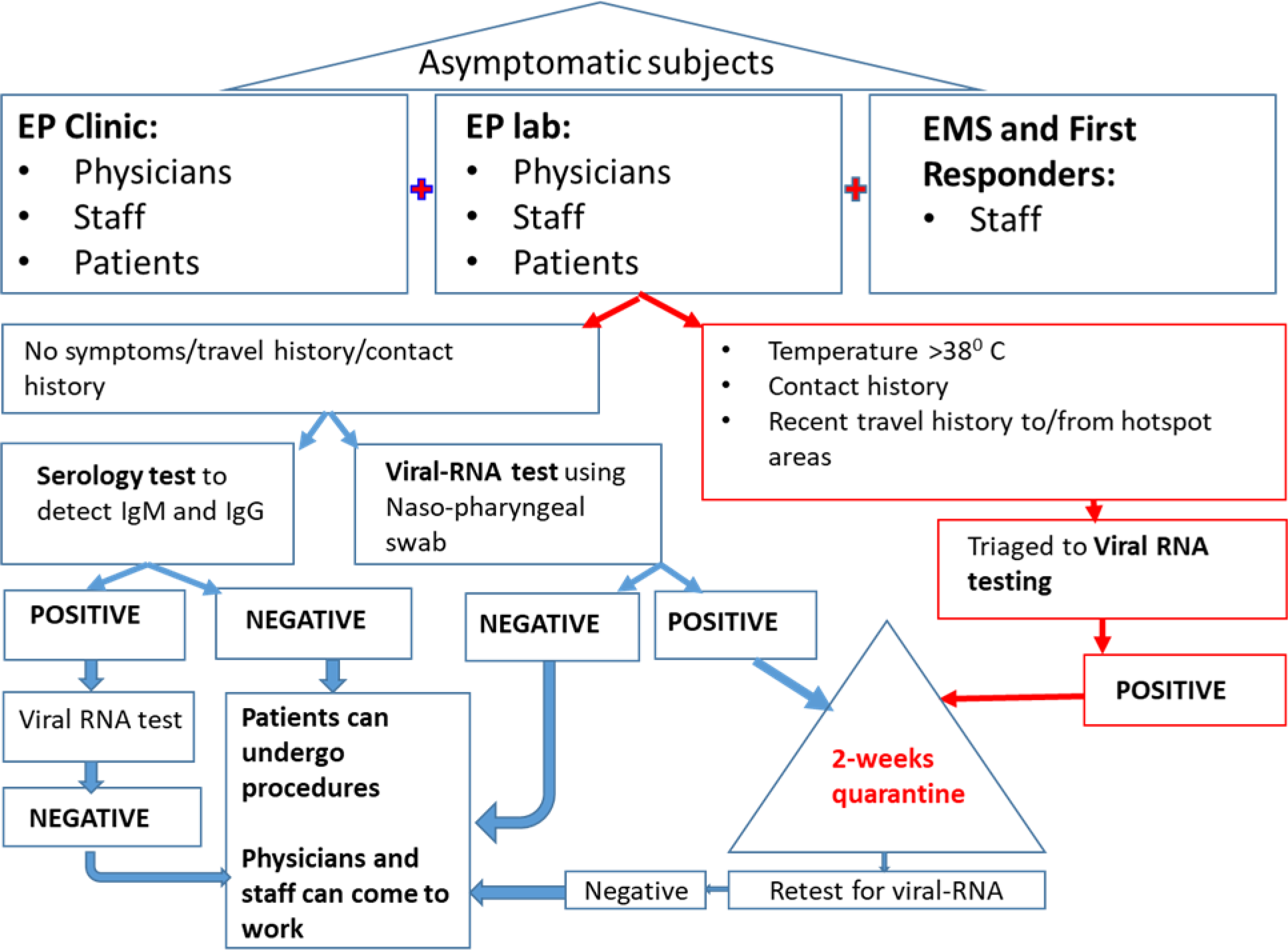
Work-flow of the study with testing and care continuum.

All patients undergoing EP procedures received general anesthesia and were intubated as necessary. The physicians and staff involved in the study, after testing negative in the viral-RNA test, interacted normally in the work environment without using social distancing and extensive protective gears. Standard precautions were taken in the lab by the anesthesia teams while intubating and extubating patients. Staff testing positive for viral-RNA on day 0 were retested on day 14 before they got back to work. Those with potential exposure to confirmed cases were retested with a swab test and contacts were traced. In 2 hospitals, personnel involved in patient care including technicians, nurses, physicians, dieticians, janitorial staff were retested at 14 -21 days using nasopharyngeal swab test for viral-RNA.

Patients undergoing EP procedures were followed up for 2 weeks after being discharged from the hospital. They were asked to measure daily temperatures and report any symptoms of COVID-19 if experienced during the 2-week period.

Patients were enrolled in our COVID-19 registry study (Clinicaltrials.Gov: NCT04352764) with an institutional IRB-approved protocol.

### Statistical analysis

Continuous data were reported as mean (standard deviation) and categorical data were presented as absolute frequencies (percentage). Descriptive analysis was performed summarizing age, gender, risk factors, presenting symptoms, and comorbidities. Individual patient characteristics were presented in a table format for patients having positive COVID-19 test result. The Student t test was used to compare continuous data and χ2 test was used to compare categorical data. All tests were 2-sided and p<0.05 was considered as statistically significant. The analysis was performed using SPSS Statistics 25.0 (IBM SPSS Inc., Chicago, IL, USA).

## Results

Starting from April 2, 2020, a total of 1670 asymptomatic subjects were tested for COVID-19 using rapid testing methods.

The Abbott ID Now kit for viral-RNA detection was validated in 46 known positive cases. Of the 46, 45 were positive on ID Now platform (sensitivity 97.8%, positive-predictive value 100%). Similarly, 58 negative specimens gave a negative result on the Abbott test (100% specificity). Serology tests were authenticated in 22 known positive and 22 negative cases (100% sensitivity and specificity). Based on our validation, we found the test kits to be reliable. No subsequent disagreements were observed during the study period.

Baseline characteristics and symptoms are presented in **Table 1**. Of the 1670 cases, 758 (45.4%) were patients attending EP clinic or lab and the remaining included caregivers of patients (n=221), EMS staff (n=219) and staff from EP clinic and lab (n=472). Majority of the physicians, hospital staff and EMS members were tested in the first week of the study period.

**Table 1:** Clinical characteristics of Population 1:

| Variables | All<br>N=1670 | Patients<br>N=758 | Healthcare<br>workers/caregivers<br>N=912 |
| --- | --- | --- | --- |
| Age | 51.94 ± 15.6 | 63.3 ± 13.0 | 42.5 ± 10.4 |
| Male | 1014 (60.7) | 545 (71.9) | 469 (51.4) |
| Female | 656 (39.3) | 213 (28.1) | 443 (48.6) |
| Returned from high-risk countries | 9 (0.5) | 9 (1.2) | 0 (0.0) |
| Close contact with anyone who traveled within the last 14 days to one of high-risk countries? | 59 (3.5) | 36 (4.7) | 23 (2.5) |
| Close contact with or cared for someone diagnosed or suspected with COVID-19 | 314 (18.8) | 35 (4.6) | 279 (30.6) |
| Chronic Lung Disease | 155 (9.3) | 89 (11.7) | 66 (7.2) |
| Diabetes | 158 (9.5) | 109 (14.4) | 49 (5.4) |
| Cardiovascular comorbidity | 352 (21.1) | 329 (43.4) | 23 (2.5) |
| Chronic Renal Disease | 36 (2.2) | 32 (4.2) | 4 (0.4) |
| Chronic Liver Disease | 17 (1.0) | 13 (1.7) | 4 (0.4) |
| Immunocompromised | 34 (2.0) | 10 (1.3) | 24 (2.6) |
| Neurological Disorder | 43 (2.6) | 21 (2.8) | 22 (2.4) |
| Pregnant female | 15 (0.9) | 0 (0.0) | 15 (1.6) |
| Current Smoker | 111 (6.6) | 84 (11.1) | 27 (3.0) |
| Former Smoker | 259 (15.5) | 154 (20.3) | 105 (11.5) |
| Atrial fibrillation | 112 (6.7) | 104 (13.7) | 8 (0.9) |
| Hypertension | 100 (6.0) | 57 (7.5) | 43 (4.7) |

A total of 64 (3.8%) cases were positive for viral-RNA at baseline. They included 31 (4.09%) patients, 26 (5.5%) EP staff, 2 (0.9%) caregivers and 5 (2.28%) EMS staff (FIGURE 2). The positive rate was 2.79% (21/753) in the month of April, 3.81% (13/341) in May and 5.2% (30/576) in June (p=0.01) (FIGURE 3). All 33 staff members testing positive on day 0 were retested for viral RNA on day 14; 32/33 (96.9%) were negative and were allowed to get back to work. One staff with positive viral-RNA on day 14 tested negative on day 21. Serology test was positive in 63/64 (98.4%) on day 14-21; 34 (53%) were positive for both IgM and IgG, 29 (45.3%) for IgG only.

**Figure 2:**
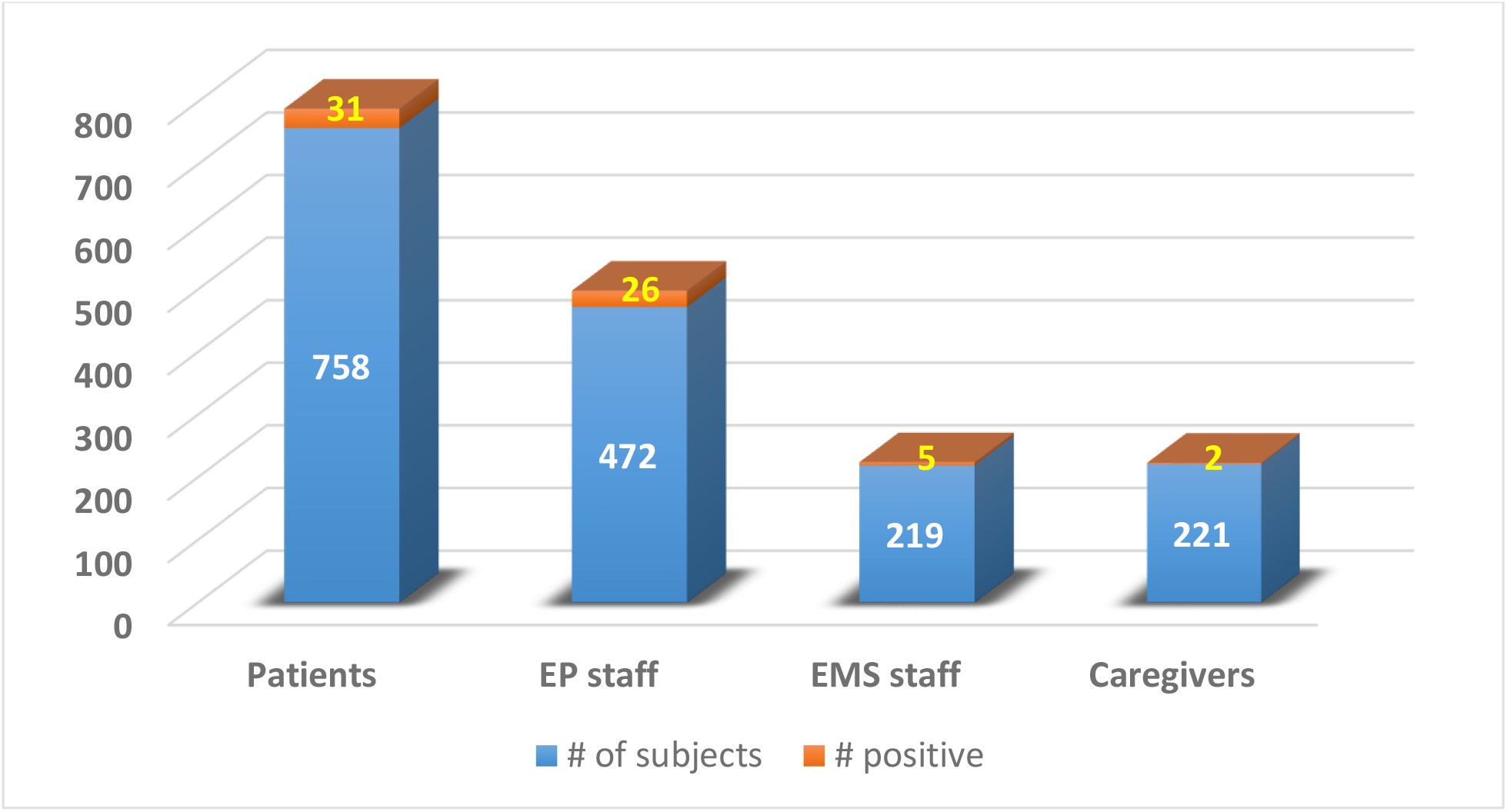
Bar diagram showing distribution of viral-RNA positive cases among the asymptomatic patient population. A total of 64 (3.8%) cases were positive for viral-RNA at baseline. They included 31 (4.09%) patients, 26 (5.5%) EP staff, 2 (0.9%) caregivers and 5 (2.28%) EMS staff

**Figure 3:**
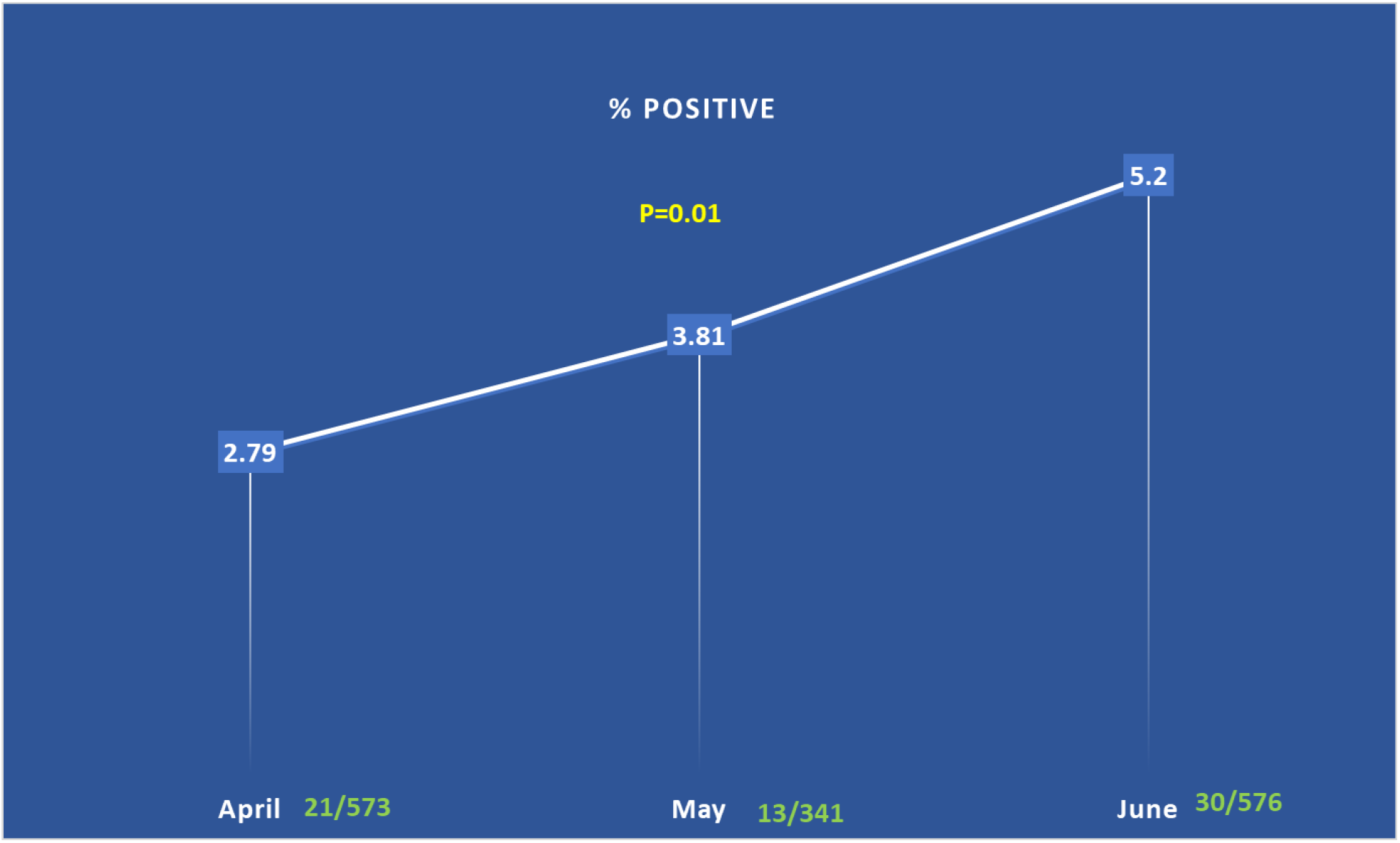
Graph showing increase in the rate of positive cases among asymptomatic individuals from April to June 2020. The positive rate was 2.79% (21/753) in the month of April, 3.81% (13/341) in May and 5.2% (30/576) in June (p=0.01)

None of the patients reported symptoms of COVID-19 infection during the hospital stay or within 2 weeks after the procedure.

### Retesting

A total of 237 staff were retested during the study period either because of potential exposure (n=67) or as part of the standard institutional practice (n=170); 6/67 (8.9%) and 0/170 tested positive. In these 6 positive cases, contact tracing revealed the possible source of infection to be family members or friends outside of the workplace. None of the coworkers of these 6 staff tested positive at that time.

## Discussion

As the continuing surge in COVID-19 cases is affecting lives world-wide and seriously challenging the reopening of institutions, businesses and borders, universal testing to identify infectious cases to limit the spread seems to be the only logical way out of this pandemic besides vaccination. However, very few places have actually implemented it into practice. Therefore, we designed this study to assess the efficacy of our universal-testing model where all staff members of the EP unit as well as all patients scheduled for EP procedures and their caregivers with no symptoms of the SARS-CoV-2 infection were tested for COVID-19 and only those testing negative were allowed to provide or receive care. Furthermore staff were retested before coming back to work if they were tested positive initially. Additionally, contact-tracing was performed to identify potential source of infection for newly positive staff members.

Our main findings were the following: 1) in asymptomatic patients and staff, the overall rate of positive cases was low (3.8%). However, it showed a significant increase in trend from the month of April to June, 2020; 2) universal testing promoted a safe environment for patients as shown by the absence of newly developed COVID-19 infections during hospital stay or immediately afterwards 3) repeat testing ensured an infection-free work environment where physicians and staff testing negative for COVID-19 could interact freely without using significantly restrictive, expensive non-conventional protective gears and difficult to implement social distancing rules and 4) the safety of workplace was further validated by identifying the possible source of infection by contact-tracing, to be from outside of the hospital in newly positive staff members. Everyone was encouraged to maintain social distancing, avoiding unnecessary public place interaction, protective masks, frequent handwashing and appropriate sanitizing outside the hospital setting. With this strategy there were no new infections amongst the patients and the EP unit could operate normally providing quality service to the patients.

After COVID-19 was declared a pandemic and a national emergency in March, 2020, social distancing, lock-down of several non-essential businesses and institutions and restrictive control measures in the healthcare settings were imposed in the USA. Several professional societies and regional authorities recommended shutting down non-emergent services and all elective procedures were placed on hold (6). Although these actions have resulted in controlling the spread of the infection to some extent, it has not only taken a devastating toll on the global economy and social fabric, but it also has ‘locked out’ other patients from receiving essential care that could have significant downstream impact on mortality and morbidity from conditions such as heart diseases, cancer and mental illness. As reported from China, many patients who had symptoms of acute myocardial infarction, presented much later to the hospital leading to significant increase in related complications, morbidity and mortality (2). Therefore, in the present context, it is critical to have an action plan that will help creating and executing a relative COVID-safe care continuum and operational abilities to continue to provide safe and high quality healthcare.

In our series, viral-RNA test was performed in all staff members, incoming patients and their caregivers and they were recommended self-quarantine for 2 weeks if positive. Thus, our screening method identified infection-free staff and patients that could safely work in close proximity without any fear of horizontal transmission of COVID-19. It not only kept the medical units functional without any hospital-acquired COVID-19, but it also saved many jobs.

The tests used to screen our subjects with no symptoms of COVID-19 revealed very low rate of positive cases. However, it was an important finding as without testing, identification of these infected asymptomatic individuals would have definitely been missed and they could have served as a source of infection for others in the workplace. Thus, by identifying and isolating them, we could prevent the spread of infection in patients and staff in the EP units.

The gradual increase in the rate of positive cases among asymptomatic individuals is an alarming finding of our study and further emphasizes the gravity of the need for universal testing to prevent the spread of this pandemic.

Our approach to control new infections illustrated the benefits of universal testing which led to a safe work environment with no hospital-acquired new COVID-19 infection in staff and patients. Furthermore, in a workplace where the staff were infection-free, with all incoming patients tested for COVID-19 for exclusion of positives, there was no need to retest all workers unless there was a reported potential exposure off work. Physicians and other staff members could interact freely in a close work-environment without using protective gears and social distancing. This model can presumably be expanded to larger populations. The current approach can also help properly triage the ongoing crisis of COVID-19.

## Study limitations

There are several limitations to the current study. Obviously, this is a small cohort from a single service line of a few hospitals. However, this provides a practical framework which can be implemented on a larger scale to restart medical services by identifying infected people in a health care environment through local testing. The tests are limited by the false positives and false negatives which can impact the care continuum. We cross verified the quality of these tests by checking them in patients with known positive and negative COVID status. The safe reboot of the healthcare activity is critically dependent on many other factors other than access to testing including – prevalence and incidence of COVID in each geographic area; hospital capacity for beds, ICUs, ventilators, personal protection equipment and above all willingness of the patients and healthcare workers to embrace elective procedures. Our study is a small beginning in a long path that needs to be laid down to create a relative COVID safe care continuum.

## Conclusion

In the current study, patient care at our EP facilities were safely carried out without any reported hospital-acquired COVID-19 infection, when prior serological and viral-RNA test were utilized to screen out all positive cases among the asymptomatic staff, patients and their caregivers. Furthermore, by assuring COVID-negative status in all healthcare workers, a safe working environment was created where all could function freely without the apprehension and risk of contracting new infection. The success of our model in creating a safe workplace by testing all individuals including those without any symptoms of COVID-19, within the confines of a specific work environment sets the example that could be utilized to keep other populated places such as offices, schools and hospitals etc. safe and open.

## Data Availability

Deidentified data will be available only with the permission of the corresponding author and for research purpose only

